# Patient perspectives on healthcare at the time of COVID-19 and suggestions for care redesign after the pandemic: a qualitative study in all six WHO regions

**DOI:** 10.1101/2021.04.06.21254840

**Authors:** Tanja A Stamm, Yuki Seidler, Margaret R Andrews, Mohammad Eghbali, Juliet Kiguli, Valentin Ritschl, Maisa Omara, Gertraud Schaffer, Erika Mosor

## Abstract

**Objective:** The COVID-19 pandemic has triggered significant changes in healthcare. As they were mainly driven by professionals and are likely to influence healthcare in the future, it is of utmost importance to consider patients’ perspectives equally. We, therefore, explored the lived experiences of patients and patient representatives in all six World Health Organisation (WHO) regions regarding healthcare at the time of COVID-19 and extracted suggestions for care redesign after the pandemic.

**Methods:** We conducted semi-structured interviews until saturation. Thematic analysis followed a modified form of meaning condensation. We established rigour by transcript checking, inter-coder agreement, quote variation and standardised reporting.

**Results:** Disadvantaged people experienced an unprecedented inequity in healthcare due to the pandemic. The main reasons were the reduction in public care services and limited access to information, transportation, technology and income. Stigmatisation from COVID-19 differed between cultural contexts and ranged from none to feeling *“ashamed”* and *“totally bashed”*. Participants experienced telehealth as indispensable but with limitations. These included giving *“bad news*”, such as having an eye removed because of melanoma, and the difficulty of providing end-of-life care over the phone. Patient representatives redefined their role and became indispensable influencers throughout the pandemic and beyond.

**Conclusion:** We reached out to individuals with a diversity of perspectives, including minorities and marginalised populations. A systematic exclusion of people with limited technology access increases inequity in healthcare and biases research findings. Since preferences and personal meanings drive behaviour and could be foundations for targeted interventions, they must be considered in all groups of people to increase society’s resilience as a whole.

## Introduction

While the COVID-19 pandemic has already triggered significant healthcare changes [1-3], it continues to challenge health professionals, systems of care and authorities. As healthcare changes were almost exclusively driven by professionals and are potentially super-influential on how care is going to be delivered in the future, it is of utmost importance to ensure that patients’ perspectives are heard and taken into account [4]. A large body of literature shows that patients’ preferences differ substantially from those of professionals. Both perspectives are essential in designing healthcare to generate value for both individuals and society [5, 6]. Patient-centricity requires that we tailor interventions to patients’ needs, uncover and understand the motivation for behaviour, and measure outcomes that matter to patients [7, 8]. In particular, patients with chronic conditions could be a rich resource for optimising healthcare as they require continuous care [9]. Furthermore, insights from different countries could show a range of experiences with disparate strategies and practices. Besides the negative impact, the crisis could thus generate opportunities for future healthcare redesign.

Patient representatives are key informants with oversight. They own a rich understanding of larger patient communities’ preferences and views, advise other patients, and deliver peer support [10]. They are close to individual patients in their local contexts [11], and many are patients themselves. Although recommendations for post-COVID-19 care designs already exist [12-14], they do not sufficiently capture and represent patients’ and their representatives’ bottom-up perspectives at a local, micro-level. We, therefore, interviewed patients and patient representatives from different chronic disease areas in all six WHO regions (America, Africa, Eastern-Mediterranean, Europe, South-East Asia and Western-Pacific) on their lived experiences regarding healthcare at the time of COVID-19 and identified key issues that could feed into future healthcare redesign.

## Methods

### Study design and participants

We conducted a qualitative study based on semi-structured interviews. We recruited patient representatives, most of them being patients themselves, over the age of 18, from different chronic disease areas through purposive snowball sampling [15] by contacting patient organisations and care providers. Chronic diseases or conditions were defined as any non-acute health problems that require continuous care. We approached eligible individuals by e-mail or telephone and explained the objectives and procedures of the study. We set up a separate interview appointment with individuals who consented to participate. We transcribed and analysed the interviews parallel to subsequent data collection to achieve maximum diversity in recruitment regarding gender, age, type of chronic conditions, country of origin and rural or urban setting. We excluded only persons with severe hearing impairment. Recruitment stopped when data saturation was met, and no new codes were identified in at least five subsequent interviews [16, 17]. The study was approved by the Ethics Committee of the Medical University of Vienna (EK Number 1388/2020).

### Procedures

The interview guide was co-developed with a patient research partner (GS) and pilot tested[18]. After agreeing on the interview procedures, EM, MO, VR, YS, and TS conducted interviews in Arabic, English, French, German, Italian, and Japanese between November 2020 and January 2021. Interviews were audio-recorded with or without video images depending on consent. Basic background information such as age, gender, disease area and years of working as a representative was collected. The interview questions focused on the challenges, perspectives, needs, preferences and prospects of care concerning the COVID-19 pandemic from participants’ views (Supplemental Table A).

### Data analysis

Thematic analysis followed a modified form of meaning condensation[19, 20] and was facilitated using ATLAS.ti [8.0] [21]. We applied the following steps: i) familiarising with data by reading through the transcripts; ii) highlighting ‘meaning units’ defined as sections of transcripts considered as relevant for our research topic; iii) assigning preliminary codes to the meaning units; iv) finalising a codebook by consolidating and revising preliminary codes; v) applying the codebook to all transcripts by still allowing adding new codes if needed; and vi) grouping codes under main themes.

### Rigour of the qualitative analysis

We checked transcripts against the recording to ensure accuracy [22]. Three authors experienced in qualitative analysis (EM, YS and TS) agreed on the meaning units. EM and YS independently coded the first six transcripts and calculated inter-coder agreement based on the percentage of overlapping codes [23]. We established transferability by collecting meaningful quotes from a wide range of participants focusing on different chronic diseases, diverse geographical and cultural backgrounds and various contexts. We reflected on our roles and perceptions influenced by our previous engagements in research that involved chronically ill patients. Finally, we reported the results according to the Consolidated Criteria for Reporting Qualitative Research Checklist[24] (Supplemental Table B).

## Results

Data saturation (Supplemental Table C) was reached after 34 interviews with individuals from 24 countries (Table 1). Nineteen participants (58%) were patients with a chronic disease themselves. In total, we identified 59 codes which we then grouped under four main themes (Table 2; Supplemental Tables D and E). Inter-coder agreement ranged between 80 and 83% (Supplemental Table F).

**Table 1.** Characteristics of the participants.

| No | Age group (years) | Years working as a representative | Sex | Country | Disease area | Diagnosed her-/himself |
| --- | --- | --- | --- | --- | --- | --- |
| 1 | 40-49 | 14 | Female | Argentina | Diseases of the musculoskeletal system and connective tissue |  |
| 2 | 60-69 | 36 | Male | Australia | Certain infectious and parasitic diseases | x |
| 3 | 40-49 | 0.4 | Female | Australia | Neoplasms | x |
| 4 | 60-69 | 26 | Female | Austria | Diseases of the musculoskeletal system and connective tissue | x |
| 5 | 50-59 | 34 | Female | Austria | Diseases of the musculoskeletal system and connective tissue | x |
| 6 | 40-49 | 16 | Female | Austria | Frailty and cognitive impairment due to ageing |  |
| 7 | 40-49 | 21 | Female | Austria | Diseases of the digestive system | x |
| 8 | 30-39 | 6 | Female | Austria | Diseases of the musculoskeletal system and connective tissue | x |
| 9 | 50-59 | 10 | Male | Azerbaijan | Chronic disease, general |  |
| 10 | 60-69 | 20 | Female | Canada | Diseases of the musculoskeletal system and connective tissue | x |
| 11 | 60-69 | 7 | Female | Canada | Diseases of the musculoskeletal system and connective tissue | x |
| 12 | 60-69 | 25 | Male | China | Chronic disease, general |  |
| 13 | 50-59 | 12 | Female | Cyprus | Diseases of the musculoskeletal system and connective tissue | x |
| 14 | 50-59 | 20 | Female | Denmark | Diseases of the musculoskeletal system and connective tissue | x |
| 15 | 20-29 | 8 | Male | Egypt | Certain infectious and parasitic diseases |  |
| 16 | 60-69 | 31 | Male | Germany | Diseases of the musculoskeletal system and connective tissue | x |
| 17 | 60-69 | 27 | Female | India | Neoplasms | x |
| 18 | 30-39 | 5 | Male | Iran | Endocrine, nutritional and metabolic diseases |  |
| 19 | 50-59 | 5 | Female | Israel | Diseases of the musculoskeletal system and connective tissue | x |
| 20 | 50-59 | 8 | Female | Italy | Diseases of the musculoskeletal system and connective tissue | x |
| 21 | 40-49 | 25 | Female | Italy | Diseases of the musculoskeletal system and connective tissue | x |
| 22 | 50-59 | 20 | Male | Japan | Diseases of the musculoskeletal system and connective tissue | x |
| 23 | 50-59 | 9 | Female | Japan | Frailty and cognitive impairment due to ageing |  |
| 24 | 70-79 | 16 | Male | Mexico | Neoplasms |  |
| 25 | 50-59 | 13 | Female | Portugal | Diseases of the musculoskeletal system and connective tissue | x |
| 26 | 40-49 | 12 | Male | Russia | Certain infectious and parasitic diseases |  |
| 27 | 40-49 | 15 | Female | Senegal | Certain infectious and parasitic diseases |  |
| 28 | 50-59 | 20 | Female | Senegal | Certain infectious and parasitic diseases |  |
| 29 | 50-59 | 28 | Female | S. Africa | Diseases of the musculoskeletal system and connective tissue |  |
| 30 | 50-59 | 24 | Female | Uganda | Endocrine, nutritional and metabolic diseases |  |
| 31 | 20-29 | 8 | Male | UK | Diseases of the musculoskeletal system and connective tissue | x |
| 32 | 50-59 | 13 | Female | UK | Diseases of the musculoskeletal system and connective tissue |  |
| 33 | 40-49 | 4 | Female | Ukraine | Certain infectious and parasitic diseases | x |
| 34 | 30-39 | 10 | Female | USA | Neoplasms | x |

**Table 2.** Main themes and related codes.

| Main themes | Codes | No.* |
| --- | --- | --- |
| Theme 1: Increasing inequity in the context of care | Overload of the health system | 1 |
|  | Reduced care and support | 2 |
|  | Test accessibility/affordability/availability/frequency/necessity | 3-7 |
|  | Restrictions accommodation and transportation | 8-9 |
|  | Fear of getting COVID-19/ infecting others/side effect medication/suboptimal care | 10-11, 13-14 |
|  | Fear (and real) drug shortages | 12 |
|  | Loneliness and mental health | 15 |
|  | Managing chronic disease in times of COVID-19 | 16 |
|  | Reduced physical functions | 17 |
|  | Reduced preventative measures | 18 |
|  | Restrictions in the basic supply | 19 |
|  | Increased solidarity, empathy and respect | 20 |
|  | More time to manage daily life and own health/to think and reflect | 21-22 |
|  | Needs for extra support | 23 |
|  | Migrants/Older adults/People with dementia/People with physical disability/Rural residents/ Socio-economically-disadvantaged | 24-29 |
|  | Disease stigma (other than COVID-19), discrimination and racism | 30 |
|  | Violence | 31 |
|  | Changes over time: Positive and negative | 59 |
| 2. Stigma and discrimination in cultural context | Stigma, COVID-19 infection/other diseases | 32 |
|  | Stigma, COVID-19 testing | 33 |
| 3. Telehealth indispensable now and in the future, but with limitations | A learning experience | 34 |
|  | Creativity | 35 |
|  | Positive statements (incl. opportunity) | 36 |
|  | Not sufficient | 37 |
|  | Technical, physical and usability barrier and accessibility | 38 |
| 4. Patient representatives as essential connectors and influencers | Creating a safe environment/Making efforts to continue care | 39, 45 |
|  | Empowering communication/Having trained and competent Healthcare providers/ Healthcare providers' communication that considers patients' COVID-related fears/Re-assuring and honest communication | 40, 42-43, 46 |
|  | Financial support continued | 41 |
|  | Information from a trusted person | 44 |
|  | Communicator and influencer | 47 |
|  | Support to others | 48 |
|  | Being challenged as representatives/advocates and reasons | 49 |
|  | Reluctant influencer in vaccination | 50 |
|  | Vaccination advantages including hope | 51 |
|  | Vaccination availability/freedom of choice/ information needed (types of information needed)/not the ultimate solution/ uncertainty | 52-56 |
|  | COVID-19 as an opportunity (other than telehealth) | 57 |
|  | Understanding both sides | 58 |
|  | Changes over time: positive and negative | 59 |

\*Code numbers correspond to Supplemental Table D. Codebook — Codes and definitions

### Theme 1: Increasing inequity in the context of care

While inequity in governance and infrastructure in care was evident between countries, the interviewees also described unprecedented disparities within their own countries. People with low self-efficacy, health literacy, education and socioeconomic status, and people with mobility limitations, migration background, older age or limited access to technologies and transportation experienced disadvantages in care. While these disparities had already existed before the pandemic, they got significantly worse due to COVID-19.

*“One of the biggest concerns always was that people were getting lost in the gaps in the* [care] *system. But now, even the more educated ones are struggling, and many more* [less-educated people] *are slipping through. I suspect there will be more long-term consequences because of that than COVID deaths*.*”* (United Kingdom, male, 20-29 years)

*“A poor woman* [with a chronic disease] *who wanted to have an abortion because she already had four children and she and her husband had lost their jobs due to the pandemic, could have gone to a private hospital and pay for it, but she had no money for that. So the child was born, an unassisted birth, and died afterwards*.*”* (Ukraine, female, 40-49 years)

One of the main reasons for increased disparities was reduced public care services due to the allocation and concentration of existing resources to COVID-19. Patients from all WHO regions were affected, but vulnerable populations in countries with already limited resources were hit the hardest. While the possibility of drug shortages “*sent a ripple of concern among our community”* (Australia, male, 60-69 years) in some countries, these shortages became a reality in others. A patient representative from Mexico explained that some “*transplant programmes* [in public hospitals] *had been shut down since March* [until December 2020]*”* and patients were sent back to dialysis, whereas private clinics almost continuously offered transplants when patients paid by themselves.

Similarly, a patient representative from Ukraine reported that when public services closed, HIV patients “*were not even welcome in private hospitals”*, even if they could pay. In Uganda, the loss of income, lack of transportation, permits required from local authorities to visit health facilities, curfews, and physical violence experience resulted in people not seeking treatment when needed. Moreover, people resorted increasingly to traditional, herbal therapies.

*“The turn up* [to the hospital] *was very, very poor, and this affected my diabetic and hypertension patients who were taking drugs regularly. I actually lost three of my clients, and I associate it with the lack of transport to come to the health facility. So, COVID prevention, the quarantine, it was so bad, the security was involved, they beat up people, and that scared the people. Apart from diabetic patients, we lost a mother who was pregnant who was supposed to have a caesarean section*.*”* (Uganda, female, 50-59 years)

Migrants with chronic diseases in Russia received treatment only from their countries of origin as many of them were not entitled to state-funded care in Russia. Due to lockdown, *“this transnational care of provision of treatment was interrupted”* (Russia, male 40-49 years). In some cases, it was even reported that the ambulance did not come if the caller had a strong foreign accent. The language barrier, weak social networks, and not having internet access were barriers to accessing information and extra support such as those provided by non-governmental organisations.

Chronically ill patients lived with multiple fears during the pandemic. While some patients were afraid of hospital infections and self-isolated from care services, others were anxious about the consequences of *“home isolation”* on physical, mental and social health (Argentina, female, 40-49 years). A chronically ill patient from the UK felt *“guilty”* for occupying care services that someone else would otherwise need for COVID-19 care (male, 20-29 years).

Despite these negative impacts, more than one-third of the interviewees also mentioned that COVID-19 has raised awareness of disadvantaged people’s situation and needs and increased the communities’ solidarity. Some interviewees suggested that health professionals should assess and address these inequities in their local contexts in the future and learn from other countries.

*“I thought that it was good to talk to your university explaining what the situation in Mexico is. It is very problematic, and we hope that, in this window of opportunity, we can change Mexico’s health system because that is what we need to come out of this with a more robust, with most equality — a system for the population especially for the most vulnerable ones which is the poor ones*.*”* (Mexico, male, 70-79 years)

### Theme 2: Stigma and discrimination in cultural context

The interviewees described social and cultural determinants of stigma, which concerned COVID-19. A patient representative from Japan explained that fear of stigma from COVID-19 prevented informal caregivers from visiting their relatives with dementia but suspected that it is different in other countries.

“*In big cities, it is less problematic, but the problem is in rural areas with very low infection rates*. [There] *the fear of being the “patient zero” is extremely high*. […] *I guess it is not like that in other foreign countries, right? It is like a hygienic mania. There are even rumours of people killing themselves of being ashamed of being “patient zero”, well this is just a rumour, but that’s how bad the stigma is. There is no way to hide the infection if you are living in a small community*. […] *The “patient zero” in our prefecture was a man who was visiting his old mother to provide home care. Later he went to his local table tennis club and caused a cluster. He was totally bashed*.*”* (Japan, female, 50-59 years)

The interviewees also discussed stigma from COVID-19 compared to other diseases. A patient representative from Senegal elaborated that it was more difficult to *“hide”* HIV medication in the time of a lockdown if many people lived together in a relatively small area. Another participant from Australia considered stigma from COVID-19 less impactful than stigma from other diseases.

*“And nobody was necessary blamed if you caught COVID. I am not saying they should have been. But look who got it. Boris Johnson and Donald Trump, and all these celebrities. Now, did they experience stigma as a result? I am not sure at all. With HIV, the stigma was quite different. I think we people with HIV now feel that we were neglected in terms of the amount of resources and interests in the community compared to COVID*.*”* (Australia, male, 60-69 years)

### Theme 3: Telehealth indispensable now and in the future, but with limitations

The provisions of various care services via the internet or telephone increased in most countries. Telehealth experiences ranged from substantial improvements of care, especially for those who live far away from the hospitals, to the need for in-person, face-to-face contacts as a definite requirement under certain conditions.

#### Telehealth being indispensable in the future of care

This theme referred to fewer efforts and time needed off from daily activities. These include paid work for medical appointments, a possibility to effectively overcome limitations of personal immobility and lack of transportation, better access to specialists who could be consulted even if physically distant, and a positive shift towards community- and home-based care. Interviewees described a steep learning curve on both sides, providers and receivers, and expressed the wish to keep these positive aspects of telehealth in the future. A patient representative from the USA with a chronic autoimmune disease described that her rheumatologist and she *“worked together to make teleconsultations ideal”*. She could then use these experiences herself when advising other patients. A participant from Austria (female, 50-59 years) with a chronic disease and physical impairment explained that *“collecting permanent medications via e-prescription in the pharmacy without having been at the physician’s office”* made her life *“much easier”* as it decreased transportation efforts.

#### Telehealth not a solution for everything

Even in situations where resources to implement telehealth were available, participants described clear limitations. Care with telehealth was, in general, not experienced as being on the same level as usual care. People reported anxiety about untreated symptoms, organ damages due to health conditions diagnosed late, increased incidences of flares, often due to medication changes by patients themselves, and increased pain due to a lack of therapies. Besides, the need for in-person, face-to-face contact when delivering *“bad news”* and making decisions that have a significant impact on patients’ lives was stressed. A patient with eye melanoma from Australia who recently had an eye removed explained that when “*patients were told this huge news about cancer or eye removal”*, face-to-face consultation and the presence of a trusted person were essential. Furthermore, while she *“loves the telehealth now”* due to her living far away and the fact that the *“doctors are always running late”*, she explained that:

*“I was supposed to have a six-month check-up in February this year. But because of COVID, they said that people could only come in if it were an emergency or something happening. So I spoke to the doctor on the phone, and she said, “Do you feel any differences?” When I said “No”, we left it. And then I went back, and it* [the eye melanoma] *was twice as size as it had been before. So, I don’t know if I had gone in February whether anything would have been different or not? Maybe not. But yeah, that will always be kind of in the back of my mind, I think, just wondering what if we caught it when it was smaller*.*”* (Australia, female, 40-49 years)

Other reasons given for problems in the widespread implementation of telecare in times of a pandemic were the unsuitability of specific interventions for distance care, the inability of some providers to offer appropriate tools for remote care and health monitoring and also the incapacity of patients to use new ways of care due to their low health literacy or financial resources.

#### Remote peer support

Remote support provided by patient representatives using digital means led to a similar discourse as telehealth. It attracted more people with broader geographical coverage since the start of the pandemic, especially people from younger age groups and limited mobility. By digital means, support could be continued during the pandemic. This virtual assistance was essential in regions with overwhelmed care providers that *“shifted resources towards working almost exclusively with COVID-19 patients”*, such as the Lombardy region in Italy, where peer support became an essential source of information for patients from different health areas (Italy, female, 40-49 years). However, digital support also had substantial limitations, and face-to-face contact was experienced as a necessity on some occasions. A patient representative from the USA described that it did not feel appropriate to conduct end-of-life conversations by telephone. Body language and visual clues were essential to establish a proper relationship.

*“All of the sudden, I was doing really serious visits with families over telehealth, like an end-of-life conversation or hospice conversation over the phone. It just looked so different, and I was scared that I wasn’t going to be able to provide the emotional support that was really needed for the patients and families through my physical presence with them. And it felt like, oh my gosh, my role is to provide support, and I am so reliant on seeing people face-to-face and following their body language*.*”* (USA, female, 30-39 years)

### Theme 4: Patient representatives as essential connectors and influencers

During the current crisis, patient representatives became essential information sources for other patients, especially when the representatives were also patients. The often-ambiguous information released by authorities, policymakers and experts added greater significance to this role. In some countries, patient representatives were recipients of downstream communications from the authorities, which enabled them to set up peer and professionally supported hotlines to take over from the overwhelmed care system. In contrast, a patient representative from Austria (female, 60-69 years) with a chronic autoimmune disease herself explained that she had not received any information regarding COVID-19 vaccinations from the health authorities. However, other patients regularly asked for her advice on this matter.

Patient representatives described that they also provided mental health support. Individuals who experienced mental health distress or suffered from loneliness turned increasingly to patient organisations. Like influencers on social media, patient representatives’ role became even more influential for informing large communities. Patient representatives passed on information to bigger groups of individuals, triggering motivation and health behaviour regarding testing and vaccination. This implies positive changes in the future care system as one patient representative in the UK commented:

*“Within the health service, there has been a recognition of the roles that the patient organisations can play in supporting patients, and patients themselves have discovered that. So, now they are beginning to see organisations like ourselves as embedded within the health system, not just as an “oh there’s an extra thing that appears”. That we actually are the part of the whole patient pathways, that they see a nurse, they see a doctor, they see a physio, they have a patient organisation. And that I think it will become more of the norm if anything that COVID has proved them*.*”* (UK, female, 50-59 years)

## Discussion

Our study uncovered four main themes. Each included a range of reflections, perspectives, and preferences meaningful for the participants in their particular contexts regarding time, setting, and personal factors. While COVID-19 measures were sometimes considered context-independent and similar in their effectiveness across countries [25], qualitative research adds to the understanding of these interventions’ meaning and their consequences for individuals. The importance of this lies in eliciting and reaching out to a diversity of perspectives, including those of minorities and marginalised or vulnerable populations. As perspectives and meanings are drivers for specific behaviour and could be the foundation for individualised and targeted interventions, they need to be appropriately addressed in all groups of people to increase society’s resilience as a whole.

While international organisations could best address inequity between different countries, healthcare providers should assess and search for individual disparities in the local context and tailor their interventions to overcome these. Otherwise, inequalities and stigma might increase existing vulnerabilities and marginalisation or create new ones. Access to care as an essential resource should be open to all individuals and a prerequisite for societies’ proper functioning. Seeing care through a lens of equity is thus necessary when redesigning services after this pandemic. A failure to do so could increase the gaps, and healthcare might become even more a good of luxury not equally accessible for all.

Digital tools and telehealth have become fundamental during the COVID-19 pandemic in an accelerated manner [26]. A generally positive attitude has been expressed. However, the lack of telehealth resources might create another disparity in healthcare within and between countries and exclude disadvantaged people from certain services [27]. Our findings also have important implications for digital survey research [28]; a systematic exclusion of people with limited technology access could lead to biased results. Even if the resources and capabilities exist use digital tools and telehealth, specific limitations must be considered by health professionals. The personal presence of health professionals and trusted people in delivering bad news with severe consequences for patients’ health was considered an essential aspect of care.

Public health interventions in a pandemic benefit from an effective bilateral, bottom-up and top-down information stream. While individuals’ perspectives are essential to tailor interventions to their needs, clear downstream information with specific channels for certain communities could best utilise patient representatives’ roles. Testing and vaccination campaigns, if organised, could also benefit from well-informed, engaged patients and patient representatives. Moreover, making perspectives, motivations and unmet needs explicit and transparent could help overcome stigmatisation and decrease social exclusion [29]. The variation and diversity elicited in our study is a strength and limitation at the same time. While we uncovered a considerable range of experiences between and within individuals and countries, several similarities emerged. Quantitative work needs to follow up to determine the frequencies of these experiences.

## Conclusion

We reached out to individuals with a diversity of perspectives, including minorities and marginalised populations. A systematic exclusion of people with limited technology access increases inequity in healthcare and biases research findings. Since preferences and personal meanings drive behaviour and could be foundations for targeted interventions, they must be considered in all groups of people to increase society’s resilience as a whole.

## Data Availability

The data generated in this qualitative study is not suitable for sharing beyond what is contained within this publication and its supplements. Further information can be obtained from the corresponding author.

## Acknowledgements

We would like to thank all people who took part in this study for sharing their valuable perspectives and the patient organisations in establishing contacts. We are grateful to the Regional Research and Training Center on Infectious Diseases (CRCF) of the National Hospital of Fann, Dakar, Senegal, for doing the interviews with participants from Senegal possible.

## Authors contributions

EM, YS and TS were responsible for the study concept and design. EM, YS, MO, VR and TS conducted the interviews. EM, YS and TS have verified the underlying data, extracted and analysed them. MA, ME, JK, EM, MO, VR, GS, YS and TS were involved in interpreting the data, writing and reviewing the manuscript. EM, YS and TS were responsible for visualising the results. GS is a patient research partner.

## Patient and public involvement

A co-author (GS) is a patient and patient representative. She participated in setting up the study concept as well as writing and reviewing the manuscript.

