## Supplement for "Patient perspectives on healthcare at the time of COVID-19 and suggestions for care redesign after the pandemic: a qualitative study in all six WHO regions"

**Supplemental Table A. Interview guideline**

| **Semi-structured key questions** |
|---|
| The aim is to gain a global picture of what has changed in healthcare due to COVID-19 from the perspective of patients and patient representatives.<br><br>1. What are the biggest burden and challenges for you and your organisation at the moment?<br>Prompt: With regard to your own health and healthcare, but also regarding the patients you are representing.<br><br>2. What are the positive experiences you had in recent months regarding your activities as a patient representative?<br>Prompts: What extra support due to COVID-19 have you received as a patient advocate from others? What exactly has been helpful?<br><br>3. How have you and the patients you represent experienced medical care provided by doctors and other healthcare professionals in the last few months?<br>Prompts: at the hospital, in the doctors' practice, at home<br><br>4. How do you and the patients you represent perceive COVID-19 testing and COVID-19 vaccination?<br><br>5. In your opinion, what has changed in the way healthcare is organised due to the pandemic?<br>Prompts: What was positive? What was negative? |
| **Final open question** |
| We now come to the end of the interview. Please share any final thoughts that are very important to you. |

**Supplemental Table B. "Consolidated criteria for reporting qualitative research (COREQ): a 32-item checklist for interviews and focus groups"**

| | Items | Question | Answer | Mentioned in |
|---|---|---|---|---|
| **Domain 1: Research team and reflexivity** | | | | |
| 1 | Interviewer | Who conducted the interview? | EM, MO, VR, YS, and TS | Methods |
| 2 | Credentials | What were the researcher's credentials? | ME: Master of Science<br>JK: PhD in Anthropology<br>EM: PhD Public Health<br>MO: Master in Dental Public Health<br>VR: Master of Occupational Therapy, Master of Science in Health Assisting Engineering<br>YS: Master of Science in Public Health<br>TS: Professor for Outcomes Research | Authors' affiliation |
| 3 | Occupation | What was their occupation at the time of the study? | TS: Professor for Outcomes Research; YS: Global Health Researcher; MA: Public Health Researcher; ME: Nursing Researcher; JK: Senior Lecturer for Community Health and Behavioural Sciences; VR: Data Analyst; MO: Public Oral Health Researcher; GS: Patient Research Partner; EM: Senior Outcomes Researcher | - |
| 4 | Gender | Was the researcher male or female? | Female: MA, JK, EM, MO, GS, YS and TS<br>Male: ME, VR | - |
| 5 | Experience and training | What experience or training did the researcher have? | All: Qualitative interviewing<br>MA, JK, EM, MO, YS and TS: Quantitative and Qualitative Methods (QMs) | Method |
| *Relationships with participants* | | | | |
| 6 | Relationship established | ….prior to study commencement? | EM: with five interviewees*<br>MO: with none<br>VR: with two interviewees*<br>YS: with none<br>TS: with two interviewees*<br>*involved in previous studies | - |
| 7 | Participant knowledge of the interviewer | What they knew about the interviewer? | As researchers in outcomes research especially in the field of chronic diseases | - |
| 8 | Interviewer characteristics | Characteristics reported about the interviewer | Experienced in QMs | Method |
| **Domain 2: Study design** | | | | |
| *Theoretical framework* | | | | |
| 9 | Methodological orientation and Theory | What methodological orientation was stated to underpin the study? | Qualitative methodology | Introduction, Method |
| *Participant selection* | | | | |
| 10 | Sampling | How were participants selected? | Purposive snowball sampling and maximum variation sampling | Method |
| 11 | Method of approach | How were participants approached? | E-mail and telephone | Method |
| 12 | Sample size | How many participants were there in the study? | 34 | Results |
| 13 | Non-participation | How many people? Reasons? | Two: Due to not feeling suitable<br>One: Due to time constraints | - |

| | Items | Question | Answer | Mentioned in |
|---|---|---|---|---|
| *Setting* | | | | |
| 14 | Setting of data collection | Where was the data collected? | At interviewees' home or workplace (via telephone) | - |
| 15 | Presence of non-participants | Was anyone else present besides the participants and the researcher? | No | - |
| 16 | Description of sample | What are the important characteristics of the sample? | Basic demographic data: Table 1 | Results, Table 1 |
| *Data collection* | | | | |
| 17 | Interview guide | Were questions, prompts, guides provided by the author? Was it pilot tested? | Yes, see supplemental table 1. Piloted tested with research partner (GS) | Method |
| 18 | Repeat interviews | Were repeat interviews carried out? How many? | No | - |
| 19 | Audio recording | Was audio or visual recording used? | Yes, of which 12 were with video | - |
| 20 | Field notes | Were field notes made during and/or after the interview? | Notes were taken during the interview and summarised afterwards | - |
| 21 | Duration | What was the duration of the interviews? | From 15 to 60 minutes — mean duration 33 with SD ± 9.12 minutes | - |
| 22 | Data saturation | Was data saturation discussed? | Recruitment stopped when data saturation was met and no new codes were identified in at least five subsequent interviews. | Method, Results, Supplemental Table C |
| 23 | Transcripts returned | Were transcripts returned to participants for comment and/or corrections? | Transcripts were not returned to the participants for comments and/or corrections. However; a patient researcher partner (GS) reviewed the data analysis to ensure that the results reflect the participants' views | - |
| **Domain 3: Analysis and findings** | | | | |
| *Data analysis* | | | | |
| 24 | Number of data coders | How many data coders coded the data? | Two, EM and YS | Method, Supplemental Table F |
| 25 | Description of the coding tree | Did authors provide a description of the coding tree? | Provided in Table 2 | Table 2 |
| 26 | Derivation of themes | Were themes identified in advance or derived from the data? | Themes were derived from the data | Method |
| 27 | Software | What software, if applicable, was used to manage the data? | Atlas.ti Version 8 | Method |
| 28 | Participant checking | Did participants provide feedback on the finding? | A patient research partner (GS) provided feedback on the findings | - |
| *Reporting* | | | | |
| 29 | Quotations presented | Were participants' quotations presented to illustrate the themes/findings? | Yes, as depicted in the results and in Supplemental Table E | Results, Supplemental Table E |
| 30 | Data and findings consistent | Was there consistency between the data presented and the findings? | Yes. We enhanced consistency and transparency by showing each step of the analysis process and the outcomes in detail | Method, |

| | Items | Question | Answer | Mentioned in |
|---|---|---|---|---|
| | | | | Results, Supplemental Table F |
| 31 | Clarity of major themes | Were major themes clearly presented in the findings? | Yes. In the main text and Table 2 | Results |
| 32 | Clarity of minor themes | Is there a description of diverse cases or discussion of minor themes? | Yes. We fully described and accounted for the exceptional cases and minor themes. Theme 2 is minor in terms of frequency but significant as a theme | Result |

**Supplemental Table C. Saturation table**

| Sequence | Transcripts | Number of new codes identified | Code numbers as provided in Supplemental Table D. Codebook |
|---|---|---|---|
| **1** | Austria, female, 50-59 years | **49 (first-round coding)** | 2, 3, 4, 5, 6, 9, 10, 11, 12, 13, 14, 15, 16, 17, 20, 21, 22, 23, 25, 26, 27, 28, 29, 31, 32, 33, 34, 35, 36, 37, 38, 39, 40, 42, 43, 44, 45, 46, 47, 48, 49, 50, 51, 53, 54, 55, 56, 57, 58 |
| **2** | Australia, male, 60-69 years | | |
| **3** | Australia, female, 40-49 years | | |
| **4** | Austria, female, 60-69 years | | |
| **5** | Japan, female, 50-59 years | | |
| **6** | South Africa, male 50-59 years | | |
| **7** | USA, female, 30-39 years | | |
| **8** | India, female, 60-69 years | 3 | 8, 18 and 19 |
| **9** | Canada, female, 60-69 years | 2 | 52 and 59 |
| **10** | Iran, male, 30-39 years | 1 | 1 |
| **11** | Egypt, male, 20-29 years | 0 | |
| **12** | Mexico, male, 70-79 years | 0 | |
| **13** | Argentina, female, 40-49 years | 1 | 7 |
| **14** | Italy, female, 40-49 years | 0 | |
| **15** | Austria, female, 40-49 years | 0 | |
| **16** | Senegal, female, 40-49 years | 1 | 30 |
| **17** | Austria, female, 30-39 years | 0 | |
| **18** | Azerbaijan, male, 50-59 years | 0 | |
| **19** | Canada, female, 60-69 years | 0 | |
| **20** | China, male, 60-69 years | 0 | |
| **21** | Russia, male, 40-49 years | 1 | 24 |
| **22** | Denmark, female, 50-59 years | 0 | |
| **23** | Italy, female, 50-59 years | 0 | |
| **24** | Japan, male, 50-59 years | 0 | |
| **25** | Senegal, female, 50-59 years | 0 | |
| **26** | UK, female, 50-59 years | 1 | 41 |
| **27** | Uganda, female, 50-59 years | 0 | |
| **28** | UK, male, 20-29 years | 0 | |
| **29** | Ukraine, female, 40-49 years | 0 | |
| **30** | Austria, female, 40-49 years | 0 | |
| **31** | Cyprus, female, 50-59 years | 0 | |
| **32** | Germany, male, 60-69 years | 0 | |
| **33** | Israel, female, 50-59 years | 0 | |
| **34** | Portugal, female, 50-59 years | 0 | |
| | **Total number of codes** | **59** | |

**Supplemental Table D. Codebook — Codes and definitions**

| No. | Codes | Code definitions: respective codes were applied when the following statements were made |
|---|---|---|
| 1 | Overload of the health system | Health system overload due to COVID-19 |
| 2 | Reduced care and support | Reduced care and support indicated by (but not limited to): difficulty in getting appointments, healthcare providers (HCPs) not ready or able to provide continuous care, reduced access due to protecting HCP from COVID-19 infection, and reduced care provided by family members due to distancing rules. |
| 3 | Test accessibility | COVID-19 testing: issues related to access |
| 4 | Test affordability | COVID-19 testing: issues related to price and affordability |
| 5 | Test availability | COVID-19 testing: issues related to availability |
| 6 | Test frequency | COVID-19 testing: issues related to frequency |
| 7 | Test necessity | COVID-19 testing: issues related to necessity |
| 8 | Restrictions accommodation | Patients' access to and utilisation of care is compromised due to reduced accommodation (e.g. hotels) possibilities around the hospitals.<br>Note: Always code with vulnerable population "rural residents" |
| 9 | Restriction transportation | Patients' access to and utilisation of care is compromised due to reduced transportation services or mobility in general. |
| 10 | Fear of getting COVID-19 | Fear of COVID-19 infection, especially hospital infection. Prime reasons for skipping routine check-ups. |
| 11 | Fear of infecting others | Fear of infecting others with COVID-19, especially in the hospital. A reason for skipping routine check-ups. |
| 12 | Fear (and real) drug shortages | Fear related to drug shortages, or real drug shortages. Includes procurement and logistical problems of routine medications needed for chronic patients. |
| 13 | Fear of side effect medication | Fear related to routine medication affecting the immune system and becoming more susceptible to COVID-19. |
| 14 | Fear of suboptimal care | Fear related to not receiving optimal care due to reduced care or change in drug regimens. |
| 15 | Loneliness and mental health | Loneliness due to isolation, social-distancing, reduced peer and family support. Also, use this code when it is indicated that the pandemic harmed patients´ mental health conditions - not only due to social isolation but hygiene obsession, depression, or aggression. |
| 16 | Managing chronic disease in times of COVID-19 | Managing chronic disease has already been a challenge in daily life before the pandemic but COVID-19 has added further challenges and complications. |
| 17 | Reduced physical functions | Patients reductions in physical functions and mobilities due to limited physical contacts (discontinuation of rehabilitation, physiotherapy, sports).<br>Note: Could be double coded with "reduced preventative measures". |

| No. | Codes | Code definitions: respective codes were applied when the following statements were made |
|---|---|---|
| 18 | Reduced preventative measures | Patients receiving minimal care, do not receive treatment, as only urgent cases are treated. |
| 19 | Restrictions in the basic supply | Limited basic supplies such as water, food and shelter due to COVID-19. |
| 20 | Increased solidarity, empathy and respect | COVID-19 raising awareness for chronically ill patients and vulnerable populations, creating solidarity among the healthcare providers, members and workers of patient organisation groups, increasing respect towards HCPs and patient organisations. |
| 21 | More time to manage daily life and own health | COVID-19 creating opportunity to have more time to manage daily life and own health due to the lockdown. |
| 22 | More time to think and reflect | COVID-19 creating opportunity to have more time to think and reflect. |
| 23 | Needs for extra support | Chronically ill patients in need of extra support and other type of support |
| 24 | Migrants | Migrants mentioned as vulnerable population or in the context of inequality |
| 25 | Older adults | Older adults mentioned as vulnerable population or in the context of inequality |
| 26 | People with dementia | People with dementia mentioned as vulnerable population or in the context of inequality |
| 27 | People with physical disability | People with disability mentioned as vulnerable population or in the context of inequality |
| 28 | Rural residents | Rural residents mentioned as vulnerable population or in the context of inequality |
| 29 | Socio-economically disadvantaged | Socio-economically disadvantaged groups mentioned as vulnerable population or in the context of inequality |
| 30 | Disease stigma (other than COVID-19), discrimination and racism | Disease-related stigma, leading to discrimination and racism |
| 31 | Violence | Violence in the specific context of COVID-19 |
| 32 | Stigma, COVID-19 infection/other diseases | Stigma related to COVID-19 infection/other diseases |
| 33 | Stigma, COVID-19 testing | Stigma related to COVID-19 testing |
| 34 | A learning experience | Telehealth mentioned as a learning experience as it is the future way of healthcare in a rather positive sense. Includes increased usabilities such as old people trying to learn how to use smartphones or computers, or reluctant organisations and hospitals gradually learning how to apply digital devices and new technologies. |
| 35 | Creativity | Telehealth being used creatively - such as reaching out to many and broader scopes of people, being able to communicate to many people at once, doing new things that have been never done before, etc.,telehealth forces the |

| No. | Codes | Code definitions: respective codes were applied when the following statements were made |
|---|---|---|
| | | healthcare providers and the patient advocate organisations to be more creative. |
| 36 | Positive statements (incl. opportunity) | Positive statements and advantages of telehealth. Includes potentials of telehealth in the future. |
| 37 | Not sufficient | Telehealth having its limits. It cannot replace everything such as human to human contact, end of life talk, reading emotional signs, check-ups that require physical contacts such as blood test, physiotherapy etc.<br>Note: Differentiate with the code "Telehealth technical, physical and usability barrier and accessibility" which is more of the hardware-related problem of telehealth. |
| 38 | Technical, physical and usability barrier and accessibility | Technical, physical, usability, and accessibility problems of telehealth. For example, not all have access to smartphone, computers and the internet. Even if they have access, not everyone can use it (excluding older adults, people with disability etc.).<br>Note: Differentiate with the code "Telehealth not sufficient" which is about telehealth not being able to replace all aspects of healthcare. |
| 39 | Creating safe environment | HCPs' efforts in creating a safe environment to continue care are appreciated and are expressed as positive experiences. |
| 40 | Empowering communication | HCPs' empowering way of communication is appreciated and is expressed as a positive experience. |
| 41 | Financial support continued | Continuous financial support to patient organisations is appreciated and is expressed as a positive experience. |
| 42 | Having trained and competent healthcare providers | Having well trained and competent HCPs especially in the time of pandemic is appreciated and is expressed as a positive experience. |
| 43 | Healthcare providers' communication that considers patients' COVID-related fears | Healthcare providers' considerate way of communication that takes patients' COVID-19 related fear and social issues (e.g. working conditions) is appreciated and is expressed as a positive experience. |
| 44 | Information from a trusted person | Information from a trusted person or trusted source is reassuring and is expressed as a positive experience. |
| 45 | Making efforts to continue care | HCPs' efforts to continue care that are interrupted or influenced by the COVID-19 pandemic are appreciated and are expressed as positive experiences.<br>Note: When patients' organisations are making efforts to continue support to their members or patients, use the code "Support to others by advocates". |
| 46 | Re-assuring and honest communication | Re-assuring and honest communication are appreciated and is expressed as a positive experience. |
| 47 | Communicator and influencer | Patient representatives pursuing important roles as communicator and influencer to patients they represent |
| 48 | Support to others | Patient representatives playing active roles in providing support to the patients they represent |

| No. | **Codes** | **Code definitions: respective codes were applied when the following statements were made** |
|---|---|---|
| 49 | Being challenged as representatives/advocates | Patient representatives challenged by lots of phone calls or not being able to answer to patients due to lack of or unclear guidelines. |
| 50 | Reluctant influencer in vaccination | Patient representatives are unsure of their roles as being influencers in the COVID-19 vaccination debate. Insecure about expressing strong opinions or giving decisive recommendations to the patients they represent. Patient representatives actively collect information but feel uncomfortable to advise others since they feel they are not experts in the field. For them vaccination is not an easy topic to talk about. |
| 51 | Vaccine advantages including hope | General and specific advantages of COVID-19 vaccination. Includes expression of the vaccine as the hope and solution. |
| 52 | Vaccine availability | Issues related to vaccine availability, procurement, and potential shortages. |
| 53 | Vaccine freedom of choice | Whether or not to be vaccinated expressed as freedom of individual choice. |
| 54 | Vaccine information needed (types of information needed) | General statements related to the lack of information related to COVID-19 vaccines as well as specific statements on types of information needed. |
| 55 | Vaccine not the ultimate solution | The vaccine alone is not going to solve all the problems. Includes statement such as other factors like overcoming the fear of COVID-19 infections, strict hygiene and sanitation measures, etc., need to continue in addition to the vaccination. |
| 56 | Vaccine uncertainty | Uncertainty expressed with regards to the COVID-19 vaccine. These include uncertainty with regards to its safety, efficacy, and side-effects for chronically ill patients who are on medication. |
| 57 | COVID-19 as an opportunity (other than telehealth) | General and specific statements made in regards to COVID-19 creating opportunity for the future healthcare system other than in the area of telehealth. These include (but not limited to): increased awareness on the importance of patient organisations, opportunity to link primary care with tertiary care, increased health literacy especially among the low socioeconomic population, increased awareness on the needs to reform healthcare that is more equitable. |
| 58 | Understanding both sides | Patient representatives holding a unique position in understanding both the challenges faced by HCPs and the patients. |
| 59 | Changes over time: positive and negative | Statements made on what has changed over time since the beginning of the COVID-19 pandemic up to the time of interview |

**Supplemental Table E. Example quotes per codes**

| Higher-level concepts | Code no. | Codes | Example quotes (country) |
|---|---|---|---|
| **1. Increasing gaps and inequity in the context of healthcare** | 1 | Overload of health system | – *Healthcare providers and physicians are too much obsessed with COVID-19 and they may refer any health issues to COVID-19 and, as a result, some diseases are missed accordingly.* (Iran, male, 30-32 years)<br>– *What we suffer the most is the workload outside the specialty to be able to sustain the health system.* (Argentine, female, 40-49 years)<br>– *Health system is very much overloaded and is not responsive to the needs of the chronic patient. That is why chronic patients are living in situation of very severe psychological stress.* (Azerbaijan, male, 50-59 years) |
| | 2 | Reduced care and support | – *Even after making an appointment with the doctor, you can expect a waiting time of a full day on site.* (China, male, 60-69 years)<br>– *The biggest burden is the destruction of care. Because my referral centre turned into a COVID unit. So, they could not manage the routine checks since the February, beginning of March. They made emergency only*. (Italy, female, 40-49 years) |
| | 3-7 | Test accessibility/ affordability/availability/ frequency/necessity | – *We have it everywhere! Everywhere, everyone can go and test if they have coronavirus. Now you need a letter from the doctor, but we have it everywhere. And people can just go. Just if you want to go, if you just suspect without symptoms, you can go and do the test. Very easy access.* (Israel, female, 50-59 years)<br>– *Access to the testing [is] non-existence. It has been a public policy since the beginning of the pandemic in Mexico not to test. It has been a real issue and obviously they did not put in place testing programmes for the population.* (Mexico, male, 70-79 years)<br>– *We are only testing those who have symptoms. But those who are not symptomatic, we are not really testing because of our economy. We limit the testing to COVID to people who have developed symptoms – the suspects.* (Uganda, female, 50-59 years) |
| | 8-9 | Restrictions, accommodation and transportation | – *It was difficult, to find a place to stay,* [it] *was difficult. You have lots places where patients can stay in Mumbai but because of COVID all the places were shut down and they were not letting people come and stay there.* (India, female, 60-69 years)<br>– *They live far away and if public transport does not work, they could not get their treatment.* (Ukraine, female, 40-49 years) |

| Higher-level concepts | Code no. | Codes | Example quotes (country) |
|---|---|---|---|
| | 10-11, 13-14 | Fear of getting COVID-19/ infecting others/side effect medication/ suboptimal care | – *People died because they were afraid to go to the hospital.* (Israel, female, 50-59 years)<br>– *A* [patient with a chronic disease] *at the moment will think twice about visiting doctors. Currently, I try to have as minimum contacts as possible with my doctor* [due to fear of getting infected with COVID-19 in the hospital]. (Austria, female, 50-59 years)<br>– *The eye hospital-thing* [appointment] *I probably could have pushed and gone in, but I thought well, lots of old people there, if I am sick I do not want to make them sick so I missed my appointment.* (Australia, female, 40-49 years)<br>– *There is always fear, and then there is this real possibility that one can no longer meet peers or be in groups with like-minded people, and the reduced offer of therapy for people who are in need. I think you have to weigh* [restriction measures] *against these things in such a crisis.* (Germany, male, 60-69 years) |
| | 12 | Fear (and real) drug shortages | – *The second stress during the second wave after autumn. It increased with deficit of the medical products and drugs in the pharmacies. And people started to buy more drugs than usual thinking that maybe this lockdown period will take longer period, maybe export and import of the drugs to the country will be delayed or postponed. Increase in price and the deficit of these drugs are making this situation let´s say worse from the perspective of the chronic patients and unpredictable.* (Azerbaijan, male, 50-59 years)<br>– *And there´s also been a great deal of lack of medicine. There is a great deal of lack of medication for these patients and you see it all over Mexico.* (Mexico, male, 70-79 years)<br>– *Before, people could get the medication for half year. They go to the hospital get the medication for half year but because of lockdown, they get medication for two weeks. They change the scheme, the doctor needed to change the medication scheme due to the lack of medicine.* (Ukraine, female, 40-49 years) |
| | 15 | Loneliness and mental health | – *There are still some patients* [who] *are extremely concerned and really, really, really anxious and almost afraid of the COVID situation in general. So they isolate themselves totally.* (Denmark, female, 50-59 years)<br>– *One more problem or burden is the loneliness. A lot of patients are at home, not seeing the relatives, not seeing the families, just you know, the husbands or the wives, not the grandmother – the wider family.* (Israel, female, 50-59 years) |
| | 16 | Managing chronic disease in times of COVID-19 | – *For some people, it is wearing.* […] *it´s just one of yet another stress that they add on to their living unless they have someone holding their hand a bit or helping them coordinate their care and they fall into a bit of a heap.* (Australia, male, 60-69 years) |

| Higher-level concepts | Code no. | Codes | Example quotes (country) |
|---|---|---|---|
| | 17 | Reduced physical functions | – *But when it comes to the training, the exercise, the physiotherapist, even in the beginning in spring, they were not allowed to go to the physiotherapist. I heard lots of people had troubles doing their normal exercises on a day-to-day base.* (Denmark, female, 50-59 years)<br>– *The patients are locking themselves in their house and their activity levels have significantly reduced. The families are really concerned about the deterioration of the cognitive and ADL* [Activities of Daily Living] *functions of the dementia patients.* (Japan, female, 50-59 years) |
| | 18 | Reduced preventive measures | – *Nobody is doing anything that is over and above what they have to do.* (Australia, female 40-49 years)<br>– *We haven´t been to the doctors or hospitals. If needed, we go to the pharmacy to pick up medicine but otherwise, we stay at home. For the routine check-up, we just tend to postpone.* (China, male, 60-69 years)<br>– *I can definitely say that prevention has gone down a lot because of the COVID.* (India, female, 60-69 years) |
| | 19 | Restrictions in the basic supply | – *So, when people came, they came by car or taxi or something. And then they will sleep in the car, they wanted to see the doctor. The streets were closed. There was nowhere they could eat food, there was nowhere they could cook food.* (India, female, 60-69 years)<br>– *We spend a lot more time on the phone to support our members and answer their questions. We have multiple requests for money that we can't cope with and we also have to listen. We participated in food distributions.* (Senegal, female, 40-49 years)<br>– *And another thing is that all the people including the districts have all taken to emergencies. These had impacts on our client. These are the challenges. Accessing utilities like food, it was not easy.* (Uganda, female, 50-59 years) |
| | 20 | Increased solidarity, empathy and respect | – *I think that they* [patients] *feel that there are actually being heard, their safety is being taken into consideration for once. Before they were having the same exact issues about really being concerned about going outside of their home and feeling like they were forced to do that but now I think a lot of the patients feel like that their needs have been met a little bit more now. They are more present for their apportionments. They can really show their medical team kind of who they are in their own home environment.* (UK, female, 50-59 years)<br>– *Healthcare seems to be just more a bit decent* [with more consideration to chronic patients] *than it used to be.* (UK, male, 20-29 years)<br>– *At the first lockdown, she* [my neighbour] *did the shopping* [for me]*. I also heard from many that they also had such nice neighbours or friends who said: no, you are a risk group, you'd better stay at home, I'll do it for you.* (Austria, female, 30-39 years) |

| Higher-level concepts | Code no. | Codes | Example quotes (country) |
|---|---|---|---|
| | 21-22 | More time to manage daily life and own health/to think and reflect | – *I lost 10 kilos* [laugh]. *Because I have the same timing for lunch and I´ve been more adherent to the therapy.* (Italy, female, 50-59 years) |
| | 23 | Needs of extra support | – *There are also some of us who are now so disabled that they are in a wheelchair or need a "personal" assistant and they just said that it annoys them that the accompanying person is not allowed to go to the doctor. So, they have to be picked up at the hospital entrance by someone else and somehow manage alone and I find that honestly also difficult.* (Austria, female, 30-39 years) |
| | 24-29 | Migrants/Older adults/People with dementia/People with physical disability/Rural residents/ Socio-economically-disadvantaged | – *More frequent use of digital technologies has significantly increased the gaps between those patients who can use this technology and those who cannot.* (Japan, female, 50-59 years)<br>– *If you have state insurance, you have free access to testing for COVID and antibodies.* […] *but lots of migrants* [with chronic disease] *even from eligible countries they don´t have that. In that case, they can go to private clinics and get it for a fee.* (Russia, male, 40-49 years) |
| | 30 | Disease stigma (other than COVID-19), discrimination and racism | – *There were few cases of discrimination. Human activists reported and also hotlines for migrants* [reported that]*, when you call the ambulance and if you have a strong accent, they drop the phone call and the ambulance just does not show up. Stigma also leads to leaving their disease status to the last worst moment. This is the default healthcare strategy of the migrant population.* (Russia, male, 40-49 years)<br>– *Some people* [whose treatment got discontinued do to COVID-19] *went to private hospitals. But you know people with HIV is not really welcomed in private hospitals. Not all of them accept people living with HIV.* (Ukraine, female, 40-49 years) |
| | 31 | Violence | – *The stress level* [has increased] *and the tolerance have dropped massively, especially in care and nursing. Colleagues very quickly feel personally attacked* [...]*, and reflection no longer takes place in the same way. Since the needs of the older adults cannot be met, they often react very harshly, very demanding, and aggressive. This is sometimes no longer bearable and leads to staff withdrawing, no longer approaching patients as openly or reacting inadequately themselves.* (Austria, female, 40-49 years)<br>– *The quarantine, it was so bad, the security was involved, they beat up people and that scared the people* [to come to hospital to seek care for their chronic conditions]. (Uganda, female, 50-59 years) |
| | 59 | Changes over time: Positive and negative | – *Additionally, we saw fewer patients with mild autoimmune diseases, but over time we saw our patients again with severe complications from their underlying diseases.* (Argentina, female, 40-49 years) |

| Higher-level concepts | Code no. | Codes | Example quotes (country) |
|---|---|---|---|
| **2. Stigma and cultural context** | 32 | Stigma, COVID-infection/other diseases | – *There is general stigma for getting COVID, not in the medical community but in general, yes. We know the people who get ill and recover, in the medical community now. But in the general public, in the households etc., […] we had [home visits] for [suspected] COVID patients by very special team coming with very special combination of staff. This was making people very stigmatised*. (Azerbaijan, male, 50-59 years)<br>– *There is a great fear of COVID-19, and here at the hospital, everyone knows that there are COVID-19 patients. People are afraid of me getting COVID-19 because I work at the hospital, so they run away from me. When I go home from work, I go straight home because people are scared. I had the same experience with Ebola at the time [epidemic in West Africa from 2013 to 2016]. I am used to it.* (Senegal, female, 50-59 years) |
| | 33 | Stigma, COVID-testing | – *One of the reasons for the low number of testing could be related to the fact that since COVID-19 infection rate is very low in Japan, people do not want other people to know that they are positive. Therefore, people are afraid of the stigma, which is of course problematic.* (Japan, male, 50-59 years) |
| **3. Telehealth indispensable now and in the future, but with limitations** | 34 | A learning experience | – *Virtuality has become an effective tool that always adds to face-to-face care. We realised that face-to-face is necessary but not essential. Patients have also learned to communicate by other means and that is very positive.* (Argentina, female, 40-49 years)<br>– *We have had a progress in telemedicine - let's say so - whether it remains will be seen. But at least I've had a taste of telemedicine.* (Austria, female, 40-49 years)<br>– *You have learned a little bit more, especially in recent time, where it is also important to use virtual media and to exchange ideas and to get further training. Overall, it is positive that we are gaining more experience with this because this is also something that will become more important in the future and which also makes sense in certain contexts.* (Germany, male, 60-69 years) |
| | 35 | Creativity | – *We have become creative in our effort to continue our activities. Especially members who live in the countryside and cannot come to Tokyo to join these kinds of workshops* [held online] *were very happy. They could get connected to members from all over Japan.* (Japan, female, 50-59 years) |
| | 36 | Positive statements (incl. opportunity) | – *I think many things get better like online prescriptions, online visits. In our private hospitals, yes. Some doctors made online clinics, and those are successful as patients´ fear hospitals, and doctors can see the patients not physically but online. So, it was a great success, online clinics in private sector.* (Egypt, male, 20-29 years)<br>– *We think it is a lot of opportunities. And the future will certainly move on to the digital solutions, and telemedicine and remote. This would be for sure* […]. *I think it will be very helpful also for patients because having that care solution will prevent them having to travel long time. It is a great opportunity that came from the pandemic and we will need to further develop it and help to adapt it for everyone. In the future, you will have more digital solutions* |

| Higher-level concepts | Code no. | Codes | Example quotes (country) |
| --- | --- | --- | --- |
| | | | *and we need to know how to go forward with the healthcare providers and healthcare service*. (Portugal, female, 50-59)<br>– *So many of our patients live very far. Those who have no prognosis and are not really having too many issues after treatment, are we really going to make them drive 3 to 4 hours just for like a follow-up visit? One thing that I think was really positive was that we were sort of forced to think about in a good way how we can use it to our benefit and how do we optimise the telehealth technologies to patients and physicians feeling comfortable, being able to do something over the phone so the patients don´t have to drive or they don´t have to expose themselves again whether it is COVID or regular environment.* (US, female, 30-39 years)<br>– *For me personally, the biggest advantage is processing prescriptions digitally* […]. *In the past, I had to take my time off from work [for getting prescriptions and medicines].* (Austria, female, 40-49 years)<br>– *Getting prescriptions works better now because I used to send my husband there to get them, so that I don't have to go in myself just to get a prescription and then catch something* [ex. COVID-19]. *Now you can just do it over the phone or by e-mail, that's fine. They could have done that earlier. I think it's actually fine when you have appointments "online", because then you don't have all the travel time, which I think is better. I feel better because I find the travel exhausting. I'm actually quite happy that I only have to sit down in front of the computer. For me yes, I would like to see telemedicine expand. I find it not bad as an "add-on".* (Austria, female, 30-39 years)<br>– *I could talk to more people at the same time* [using Zoom]. *I am getting to know people even better* […] *and can talk* [to people] *anywhere in Canada. I find it very therapeutic.* [Geographical] *barriers are gone.* (Canada, female, 60-69 years)<br>– *I think it would be very nice with the teleconsultation. It can be difficult to know which patient needs it or not. I sometimes only see my consultation* [doctor] *only five minutes or something and it is waste of time if there is nothing to talk about. The form every patient needs to fill out before consultations can now be filled out electronically at home. You can do it from your cell-phone or computer before seeing the consultant. That´s also a different new thing which will also be there after this COVID situation. So that´s also a very good thing*. (Denmark, female, 50-59 years)<br>– *Some of the healthcare centres or organisations provide online and remote services to the patients with less severe problems such as voice or video calls. This can help reduce referrals to the medical centres.* (Iran, male, 30-39 years) |
| | 37 | (Telehealth) not sufficient | – *Of course, it* [telehealth] *doesn't replace "face-to-face" care and everything. I know that there are many people who are sceptical and I mean, it just does not replace everything.* (Austria, female, 30-39 years)<br>– *I still have to go into the hospital and have my blood taken and the doctor will then communicate by telehealth about what the blood showed them. That´s OK and sometimes that worked but there were number of circumstances with my co-morbidities and I have quite a lot, where doctors, if the doctors did not look at me, I did not really feel like I was getting an adequate attention to the problem. All I am saying is: it´s not perfect* [laugh]. |

| Higher-level concepts | Code no. | Codes | Example quotes (country) |
|---|---|---|---|
| | | | *Telehealth is not perfect. I also think doctors pick up a lot when they are face-to-face with a person that they won´t on telehealth. I got my doctor ring me up they did not even see me face-to-face which is sort of not very good either cause you can pick up certain visual clues from when they are face-to-face. I think a lot of people didn´t have their emotional health and well-being looked after particularly well because telehealth is not or adequate at picking up the emotional grooves.* (Australia, male, 60-69 years)<br>– *It* [tele-consultation] *can be difficult to know which patient needs it or not. You have to know which patient want to do this or not, and be sure that you don´t lose anybody.* (Denmark, female, 50-59 years)<br>– *The opportunities to use digital technologies have significantly increased and this has increased the gaps between those patients who can use this technology and those who cannot. Because of the digital technology, there is an overall assumption that patients with chronic diseases or disabilities should be OK being locked in at home. This perception discourages patients to go out and exacerbates the already limited social participation of the chronically ill patients. Face-to-face communication is still important for some patients. Some patients go to the hospital because they want direct communication* [maybe more than the treatment itself]. (Japan, male, 50-59 years)<br>– *Families of dementia patients are very sensitive to tiny reactions of the patients and are always concerned if the patients are reacting to their voices or can recognise them. The spirits are heightened even with such small signs but if the visit is through videos or with doors in-between, these are difficult. Especially for patients with an advanced level of dementia. If the families cannot visit for a long time, some patients do not recognise them anymore.* (Japan, female, 50-59 years)<br>– *It* [telehealth] *is only a solution for* [individual] *case. You need to go case-by-case.* (Portugal, female, 50-59 years) |
| | 38 | Technical, physical and usability barrier and accessibility | – *What is certainly happening is that quite a few fall through the grate, where I always ask myself the question a bit, it depends on the age group and the access to the new media — electronic media, because someone at 70 who has nothing to do with smartphones and the Internet, how will s/he know that s/he can actually call the hospital and come there, sometimes too much, I think.* (Austria, female, 40-49 years)<br>– *Telehealth sometimes does not work. You know it just completely in the middle of the session cuts out, when you are trying to talk and that happened to a lot of my friends. I get a bit exacerbated by, cause you know, it is all very well to believe that the Zoom that we are on, the net that we are on is perfect but it is not.* (Australia, male, 60-69 years)<br>– *At the moment* [telemedicine is] *not working much. Why? Because the majority of chronic patients are old people and in Azerbaijan the people with certain age, older age, let´s say after 50 etc. differently from myself, for example, I am using computer but most people are not skilled enough to use the internet and the modern technical digital. They are not skilled in that very much. Exactly the same I can talk about the medical doctors of this age also. Young medical doctors they are OK to work online etc., but older doctors not at all. So, I would say maybe the capacity of telemedicine covers 5-10% of real needs at the moment.* (Azerbaijan, male, 50-59 years) |

| Higher-level concepts | Code no. | Codes | Example quotes (country) |
|---|---|---|---|
| | | | – *But there are many patients who shy away from that. Especially the older ones. There are also real aversions. We have also tried to train the people or offered training to equip them technically. I have experienced that especially the older ones say "No, I'm not familiar with that at all, I can't get involved in it, that won't work and I don't want that." So, these personal contacts and group meetings simply broke apart.* (Germany, male, 60-69 years)<br>– *The only concern I have is that, yes, we can keep it for the future as a good practice but we have to consider that teleconsultations via Webex, Zoom, Skype, WhatsApp are not secure. Exchange of very sensitive data. And also, the reimbursement of procedures must be clear. And we also need to teach both clinicians and patients, how to better use this, this option.* (Italy, female, 40-49 years) |
| **4. Patient representatives as essential connectors and influencers** | 39, 45 | Creating secure environment/Making efforts to continue care | – *And if you have volunteers who want to do activities face-to-face we are allowed to do that. But we had a whole tool for them, we made available two guidelines and also posters where they are informed on how to have physical face-to-face meeting in a secure way. Actually, we hear from a whole lot of patients coming to different activities that they feel secure and they feel that it is very nice to come because they have been sorted out and it is very, very clear in how we do it.* (Denmark, female, 50-59 years)<br>– *So there is a continual feedback with emails and phone calls and trying to reassure people that we are doing everything in our practice to keep ourselves and them save in this COVID environment.* (South Africa, female, 50-59 years) |
| | 40, 42-43, 46 | Empowering communication/Having trained and competent healthcare providers /Healthcare providers' communication that considers patients' COVID-related fears/Reassuring and honest communication | – [The assistant] *called all patients to make sure that everyone gets a check-up appointment and called every one of them again to explain how these [physical] appointments are going to take place.* (Austria, female, 40-49 years)<br>– *She* [my doctor] *was reaching out at the beginning of the pandemic asking me how I´m doing, asking about what I am thinking to do about the medication and kind of coming up with something like an emergency plan. Talking me through the information that she had, information she knew. If the things are bad what are the things I can do in the meantime and maybe I have to work from home but she* [the doctor] *would really help me with that decision. So, I felt like that I was not making that decision by myself. I feel like it has been much more patient-centred. She is really taking into consideration all these pieces of COVID that might affect my life both as a patient and as a practitioner. So that was something I really felt supported by her to help me get through the pandemic in terms of my work decisions anything like that, so I felt very, very supported by her.* (USA, female, 30-39 years) |
| | 44 | Information from trusted person | – *I would personally feel the safest if my family doctor (GP) would do that [adjusting treatment regimen to adapt to the COVID-19 situation] for me. Because I simply have confidence in him. We just have to clarify whether it is going to be with weaker doses or whether that is going to fit me or it is going to be too strong. I would just clarify that in advance with him. I would feel the most comfortable with [making decision with] him.* (Austria, female, 50-59 years) |

| Higher-level concepts | Code no. | Codes | Example quotes (country) |
| --- | --- | --- | --- |
| | 41 | Financial support continued | – *Due to the government´s "dormant deposit system" [a system that utilises dormant bank savings to be used for NGOs/NPOs activities], financial support to my associations have increased. Many different kinds of funding schemes have been initiated due to COVID-19.* (Japan, male, 50-59 years)<br>– *There were actually individual donors again because of the work we were doing and being very accessible for the patient they really begun to value us so we did have lots more individual donations because people wanted to support us to keep us going.* (UK, female, 50-59 years) |
| | 47 | Communicator and influencer | – *We do that a lot over the phone. Lots of people calling me asking information about the hospital, how to get the treatment and also, they ask a lot for psychological support and we also have a* psychologist [at our patient organisation]. *She offers her services for free. There have been a lot of contact over the telephone.* (Cyprus, female, 50-59 years)<br>– *It is also our responsibility to be telling the patient that something like this was unexpected, we have never seen it our life-cycle and also the government was not prepared for something like this. So, whatever the fuss has been made is the learning process. We start with all that. I am trying to reason out with them, I am trying to explain to them to remember how the situation was. By the end of the call, by the end of the meeting their attitude has changed. And they were like you know, "Thank you for spending so much time with us, thank you for explaining to us. And thank you for reassuring us that we can always come back to you and talk to you" and all that. So, I think it is the role of the patient advocates to be with the patients at this time.* (India, female, 60-69 years)<br>– *We already have peer-support line a number where people can call and get some support, peer support. We developed further our digital communication by email, digital newsletter. Four months ago we have issued a newsletter about vaccination, about the vaccine and pneumonia, pneumonia vaccine but also trying to inform the patients about what type of vaccines are available and also to recommend to talk to their doctors and health professionals about the vaccine to have their doubts clarified. To keep an eye on the vaccination plan. Because they are always very suspicious and they don´t know enough around vaccine. The new vaccine, there is not enough information. Therefore, we are all waiting for some recommendations or scientific guidance from the rheumatologist so that will help for sure if is safe or not.* (Portugal, female, 50-59 years) |
| | 48 | Support to others | – *It´s OK if the hospitals didn´t listen to us or the policymakers don´t listen to us, it doesn´t matter. As long as at the end of the day the patients are happy, which means we have shared with them, and we have explained things to them. I think that is important because happy patient is a better person.* (India, female, 60-69 years)<br>– *They have been a great drop in consultations. So, we have to establish a way to communicate with patients, trying to give them, first of all hope, "Hold on there, stay put, try to don´t get COVID-19, so if we can help you [we will]." A lot of patients were in haemodialysis. We [visited] those haemodialysis centres and obviously this with our resources.* (Mexico, male, 70-79 years) |

| Higher-level concepts | Code no. | Codes | Example quotes (country) |
|---|---|---|---|
| | | | – *With the coronavirus, we have adapted, we have done a lot by telephone to contact our members and answer their questions. If we learn that a person is sick, we go and pick them up and take them to the hospital here.* (Senegal, female, 40-49 years) |
| | 49 | Being challenged as representatives/ advocated and reasons | – *People are flooded with information and when they ask, who gives them the right answer and the right answer doesn't really exist. People come to us, ask specific questions such as "Is this hospital open? Can I go there? I have my follow-up appointment next week. Where should I report?" and so on? Most of the time I just keep it open. It's* [the answer] *just still open. We urgently need some kind of official statement now. Of course, they* [the authorities] *also wanted to make sure, because there was no scientific data, but at least a little bit of a guideline that we can refer to.* (Austria, female, 40-49 years)<br>– *I think one of the biggest burdens is that we lost contact with our members. We can´t have any events. So, all events are virtual. And so also funding. We have lost a lot of funding because we don´t do events. We need to justify what the money is used for. So, if there is no event, we have no funding from the government.* (Cyprus, female, 50-59 years)<br>– *I would lie if I mentioned anything positive about COVID-19. Before the pandemic, we had funds, lots of funds from lots of people and businessmen to help us and our mobile care services.* (Egypt, male, 20-29 years)<br>– *So, this for us has become very worrisome also because we had to tell the patient "Hey, your hope for a transplant will be delayed for an unknown period of time". And this is also leading to the great percentage of them dying.* (Mexico, male, 70-79 years)<br>– *We were pushed to find out how we communicate with the members and even with other stakeholders that we are trying to fight for the access to medication in this time. And all these kinds of meetings there are all virtual, [we are] not really ready for that. Everyone is getting a little bit tired of the digital demand. There is not enough time to deal with the computer and telephone calls and emails and the rest.* (Portugal, female, 50-59 years)<br>– *As associations, we haven't received anything, even though we are in the care sites and we are doing a lot of work, especially for the psycho-social support of our members. But here it's as if the Ministry didn't recognise our role, whereas we do it every day. On the financial level, we have received nothing and that is very negative. We have accumulated a great deal of experience in supporting PLWHIV and our association is effective, but this is not recognised here.* (Senegal, female, 40-49 years)<br>– *I spend a lot of time for patients who are not in good health, especially those with co-infections, with hepatitis, but also patients with depression. It is difficult to follow them at the moment and they have a lot of needs. For patients who are hospitalised, we also support them, with prescription purchases, help to pay for hospitalisation and biomedical tests. Often patients don't have companions, so I have to play this role. There's an overload of work at the moment, we're under a lot of pressure.* (Senegal, female, 50-59 years) |

| Higher-level concepts | Code no. | Codes | Example quotes (country) |
|---|---|---|---|
| | | | – *Apart from the corona, there is the poor psychosocial support. People all around are worried. Our patients, persons with chronic illnesses, are the ones who are vulnerable. Patients with chronic illness become so scared. This new style of Coronavirus has really affected these people. There have not been a lot of outreach because they have not been a lot of attention and funding for the activity to reach out to clients. Outreach, no funding, no facilitation for that.* (Uganda, female, 50-59 years)<br>– *The biggest issue in the fight at the beginning of the pandemic was the lack of clarity around who with rheumatoid arthritis or juvenile idiopathic arthritis should be classified as extremely vulnerable and needing to be shielded. There was lots of misinformation and incorrect information going out from primary care and then secondary care hospitals.* (UK, female, 50-59 years)<br>– *In March alone when we first got hit with COVID here in UK, we had a 600% increase of calls to our helpline. That´s just telephone calls. And we got equally sort of increase on emails and you know people actually sending in emails asking similar questions so there was this sudden surge which put us under a huge amount of pressure as a small organisation. it has been a huge pressure on us as an organisation, to just manage that and try to keep up-to-date with things that moving so quickly to be able to give that information in lay terms out through our various networks.* (UK, female, 50-59 years) |
| | 50 | Reluctant influencer in vaccination | – *If people ask my opinion on vaccination, I will say according to my state of knowledge that I have from the media or from here and there. I always say that this is my personal opinion and please that they also form their own opinion.* (Austria, female, 40-49 years)<br>– *I would say that I do not need to be the person first in line. I would like to see, want to know a bit more and hear about the side effects and so on and also I would like to have my rheumatologist clarify with my specific medication whether could be a problem or not.* (Denmark, female, 50-59 years) |
| | 51 | Vaccination advantages including hope | – *I think that it´s a great opportunity. I have spoken this morning to, in another meeting with a colleague, and I said "You should promote the vaccination to say that the benefits are higher than the risks, that a possible ADR drug reactions can be managed, that you can come to a normal life only with the vaccination".* (Italy, female, 40-49 years)<br>– *What it means is that it means the same thing in all the world. That it is a great hope. The great light at the end of the tunnel.* (Mexico, male, 70-79 years)<br>– *Definitely yes* [I will encourage my patients to be vaccinated]. *The chronic ill patients, if there is no complication, they have to take the vaccine because they are the most susceptible.* (Uganda, female, 50-59 years) |
| | 52 - 56 | Vaccination availability/ Freedom of choice/ Information needed/Not | – *They are kind of saying that there are enough of them, but average people like us will have to wait and see for some time. I don't expect to get a vaccine probably until May or June because they have to do all the health professionals* |

| Higher-level concepts | Code no. | Codes | Example quotes (country) |
|---|---|---|---|
| | | the ultimate solution/Uncertainty | *and the long-term care homes, the paramedics and of course we don't have that many vaccines coming in all at once. Like all the world wants them so there's gonna be a lot of wait and see.* (Canada, female, 60-69 years)<br>– *My attitude* [to vaccination] *is not really positive because my general attitude to the situation it is more political pandemic rather than real clinical biological.* (Azerbaijan, male, 50-59 years)<br>– *They are not sure. About the safety. Not about the vaccination process, about the safety of the vaccine.* (Egypt, male, 20-29 years)<br>– *There are lot of, you know, fears of the vaccines because not much is first known about the disease, so obviously nothing is known about the vaccine, the efficacy of the vaccine, how long will it be effective, so there is a lot of question-marks about it. The safety is the biggest issue and how long would it keep you safe from the disease.* (India, female, 60-69 years*)* |
| | 57 | COVID-19 as an opportunity (other than telehealth) | – *It will become more of the norm if anything that COVID has proved them. It is validated why we need patient organisation and the important role that we can play. Not just for the patients but for the healthcare professionals as well. And be a partner in the care. I think that is going to be the change and I think that´s what the patients are beginning to see as well. That looking at us now, far more in a professional way and not just, as charities, sometimes we can be seen as oh that, you know, nice sort of "hold your hand make you feel better", not actually having a real say how things will improve and having a say on the political side of things: the advocacy as well, that we do have a voice we can be talking on behalf of the population of patients that we are representing and I think that has improved. I think people begin to see us in a different light which is good.* (UK, female, 50-56 years)<br>– *Those people who are poor might have developed more understanding to the importance of health, and understanding of health concepts. We are expecting that once we can go back and support those people, they may have better understanding to their health problems* [COVID-19 being a health educating opportunity]. [Rural] *patients in Egypt lacked knowledge and education, especially in the village. They need to learn and they learn quick, that is not a big deal but we don´t get support for* [such health] *education. Something we need to do in the next years.* (Egypt, male, 20-29 years)<br>– [The possibility of remote care] *made doctors think 'why do patients need to come to a big centre'. So now what the doctors are doing now is connecting them to local doctors. And requesting the patients to go to the local doctors and see the local doctors. So, the local medical system will also get developed. Now the patients will know where to go.* (India, female, 60-69 years)<br>– *Since people are now more careful about handwashing and measures to prevent infectious diseases, there has been a drastic reduction in the number of overall seasonal infections, such as influenza. This may reduce the risks of patients with chronic diseases from developing complications from opportunistic infections.* (Japan, male, 50-59 years) |

| Higher-level concepts | Code no. | Codes | Example quotes (country) |
|---|---|---|---|
| | | | – *Our system has been so exposed in its lack of things, in its failure of things, in the lack of investment, so exposed, that it has become a positive thing because of the window of opportunity was there with all this evidence. We are really pushing in this window of opportunity to really promote change in our health system. That, to me, is a positive thing. Because we see that now we have not only a perception that we need a health reform but it is an evidence now that a health reform is needed more than ever. So, this is to me a very positive thing.* (Mexico, male, 70-79 years)<br>– *We just hope that out of this comes out really preventive programmes for people to eat better, to exercise, to take care of their consults, to really go maintain their adherence to their treatment and that we can put them in place. It is more important to control diabetic, obesity, hypertension because this is really a pandemic we have in our country and that we need to do something about it. Whether we have COVID or we don´t have COVID.* (Mexico, male, 70-79 years)<br>– *One positive thing with the COVID-19 vaccine is that it reminded a lot of people that it was important to get the basic vaccines, especially for children. We've seen that. Even here at the hospital, mothers who are late in vaccinating their child are called on the phone for vaccine reminders, and this was rarely the case before.* (Senegal, female, 40-49 years)<br>– *Something that came out of COVID is the general hygiene of the clients, the staff, the facilities have improved in general. The water-borne diseases have reduced in the region.* (Uganda, female, 50-59 years)<br>– *We have just demonstrated the massive importance of self-management, and that's what we need to consider*. (UK, male, 20-29 years) |
| | 58 | Understanding both sides | – *I do understand how a clinical trial work and they [immunocompromised patients] were probably excluded from clinical trials as long with pregnant women but it doesn't mean that pregnant women shouldn't be able to have them* [vaccines] *or immunocompromised people.* (Canada, female, 60-69 years)<br>– *Getting those guidelines even from the professional bodies was quite difficult. I mean, they were saying "Yes, we are working on it". And I know this was an unprecedented [situation] and it was difficult to give black and white answers. But it took probably too long for the professional bodies to bring that level of information out to the general public.* (UK, female, 50-59 years) |
| | 59 | Changes over time: positive and negative | – [What has changed is that] *they have included me to an electronic record for blood tests and so on. They have certainly encouraged more people to do so because they can also view their COVID test results with it.* (Canada, female, 60-69 years)<br>– *Anxieties and uncertainties related to medication were very high but rather at the beginning of the Corona* [pandemic] *when rumours about the hydroxychloroquine treatment were widely spread*. (Germany, male, 60-69 years) |

| Higher-level concepts | Code no. | Codes | Example quotes (country) |
|---|---|---|---|
| | | | – *Lots of the people taking the Rheuma medicines had fear what if they get the coronavirus and how is it going to affect us [them] — that was fear in the beginning. Today it is the fear of vaccination.* (Israel, female, 50-59 years) |

**Supplemental Table F. Details on the six steps of the data analysis.** These included: i) familiarising with data by reading through the transcripts; ii) highlighting 'meaning units' defined as sections of transcripts considered as relevant for our research topic; iii) assigning preliminary codes to the meaning units; iv) finalising a codebook by consolidating and revising preliminary codes; v) applying the codebook to all transcripts by still giving the opportunity for adding new codes if needed; and vi) grouping codes under main themes.

| Steps | Detailed explanation of the steps |
|---|---|
| **Step 1** | EM, YS and TS read through the transcripts and checked them for accuracy |
| **Step 2** | EM, YS and TS highlighted 'meaning units' defined as sections of transcripts considered as relevant for our research topic. |
| **Step 3** | EM and YS independently coded the meaning units of six transcripts. The number of the identified codes by EM and YS was 46 and 41, respectively. |
| | EM and YS compared the results by counting the numbers of the same or similar codes used for the same segments of the transcripts. The number of overlapping codes were 37. Inter-coder agreement rate was calculated by dividing the number of overlapping codes 37 with the number of codes identified by EM: 37/46 (80%). EM and YS agreed to split and merge some of the codes and agreed on new codes. |
| | A preliminary codebook was developed with 49 codes, each with definitions. |
| | EM and YS independently coded additional three transcripts using the preliminary codebook. |
| | EM and YS compared the results by calculating the percentage overlaps of each code used for the respective segments of the transcripts. For example, if both applied the code "fear of COVID-19 infection" for all three transcripts in the more or less same segment of the transcript, this was calculated as 100%. The number of new codes identified by EM and YS were 7 and 5, respectively. The average overlap from 61 codes was calculated, which was 83%. |
| **Step 4** | After merging the codes, EM, YS and TS discussed and agreed on a codebook consisting of 55 codes from which main themes were developed. |
| **Step 5** | YS continued to code the rest of the transcripts, identified 4 additional codes and discussed and agreed on the results with EM and TS. The final code-book consisted of 59 codes. |
| **Step 6** | EM, YS and TS grouped the codes under four main themes. |
